# Psychiatric morbidities and Coping strategies in patients with different Coronavirus disease-2019 severities and chronic medical diseases: A multicenter cross-sectional study

**DOI:** 10.1101/2020.12.22.20248379

**Authors:** Hend Ibrahim Shousha, Nagwan Madbouly, Shaimaa Afify, Noha Asem, Rabab Maher, Suaad Sayed Moussa, Amr Abdelazeem, Eslam Mohamed Youssif, Khalid Yousef Harhira, Hazem Elmorsy, Hassan Elgarem, Dalia Omran, Mohamed Hassany, Bassem Elsayed, Mohamed El kassas

## Abstract

COVID-19 patients, especially those with chronic medical illnesses (CMI), may use different coping strategies, to reduce their psychological distress while facing the COVID-19 infection. The aim was to compare anxiety, depression and coping styles between patients infected with COVID-19 disease with and without CMI during the peak of COVID-19 disease in Egypt. This is a cross sectional study, that included an online survey consisting of Arabic versions of General Health Questionnaire-12, Taylor Manifest Anxiety Scale (TMAS), Beck Depression Inventory (BDI) and Brief-COPE scale. Questionnaires were distributed to adult patients with a confirmed diagnosis of SARS-CoV-2 virus infection during their quarantine in Egypt. One hundred ninety-nine patients responded to the survey, where 46.73% of them had CMI. Religion, emotional support, use of informational support and acceptance were the most used coping strategies by participants. Avoidant coping strategies were frequently used by divorced patients, home quarantined individuals, patients who developed COVID-19 related anxiety/depression and patients who didn’t receive hydroxyl-chloroquine. Approach strategies were frequently used by patients with mild COVID-19. Understanding the used coping strategies has implications for how individuals might be helped to manage their illness during the current presentation and intervene with development of serious long-term mental health conditions.

## 1. Introduction

The severe acute respiratory syndrome coronavirus 2 (SARS-CoV-2), causing coronavirus disease 2019 (COVID-19), emerged in December 2019 as a multifaceted problem. It gave rise to a global health threat, resulting in an on-going pandemic in many countries and regions of the world. During the last months, it was observed that the virus caused a wide range of clinical manifestations in all age groups. Moreover, it had a negative impact on the affected individual’s life in many aspects. It would affect his/her physical as well as the mental health. In addition, it had its adverse effect on the economic and daily life activities due to the unexpected lockdown (Li et al, 2020).

Older adults and people of any age with chronic medical illnesses (CMI) are particularly at increased risk for severe illness and affection of their daily lives (Sanyaolu et al, 2020). Studies have demonstrated that chronic disease, including cardiovascular diseases, hypertension, pulmonary diseases, and diabetes mellitus are considered as risk factors for the disease severity, poor prognosis, and mortality in COVID-19. In China, the overall case-fatality rate was elevated among patients with pre-existing CMI: 10.5% for those with cardiovascular disease, 7.3% for diabetes, 6.3% for chronic respiratory disease, 6.0% for hypertension, and 5.6% for cancer (Wu & McGoogan, 2020). However, the definite reasons for the association between these comorbidities and the disease severity and mortality risk of COVID-19 have not been yet identified (Güler & Öztürk, 2020).

Coping refers to the use of psychological patterns to manage the individual’s feelings, thoughts, and actions to maintain psychological and social equilibrium during different stages of ill-health and treatments (Stanisławski, 2019). Individuals under different conditions can employ passive or active coping strategies during exposure to stressors. Passive coping strategies include refusing to acknowledge the existence of stressful conditions, giving up on making efforts to overcome the stressors and thus leading to strengthening stressful feelings. Active coping strategies include accepting the existence of stressful events, finding ways to overcome stress, taking advantage of the situation by learning lessons from it and make plans for subsequent efforts (Yu et al, 2020). This suggests the need to explore whether COVID-19 patients with and without CMI used different coping styles while facing the COVID-19 infection to reduce their psychological distress. Understanding these strategies has implications for how individuals might be helped to manage their illness during the current presentation and intervene with the development of serious long-term mental health conditions in the future.

The aim of this study was to compare anxiety, depression and coping strategies between patients infected with COVID-19 disease with and without CMI during the peak of COVID-19 disease in Egypt (June-July 2020) (Worldometers.info, 2020). It was hypothesized that COVID-19 patients without CMI would have lower psychological distress and use active coping styles compared to COVID-19 patients with CMI.

## 2. Methods

### 2.1. Subjects

This is a cross-sectional study that consists of an online survey that was conducted from 7 June to 20 July 2020. The tools of measurement were implemented in a Google drive format and distributed to adult patients (≥ 18 years) with a confirmed diagnosis of SARS-CoV-2 virus infection during their quarantine, either at their homes or in hospitals (Students Hospital, 15 Mayo Smart Hospital, National Hepatology and Tropical Medicine Research Institute) in Cairo and Giza governorates, Egypt. Confirmed diagnosis was defined as having positive results of real-time reverse-transcriptase polymerase-chain-reaction (RT-PCR) assay for nasal and pharyngeal swab specimens. Participants joined the study voluntarily after giving an informed written consent.

### 2.2. Measures

#### 2.2.1. Outcome variables

##### 1. Psychological distress

The Arabic version of 12-item General Health Questionnaire (GHQ-12) (Daradkeh et al, 2001) was used as a screening device for psychological morbidity. It is a self-administered questionnaire that has been used for both population-based studies and health assessment surveys. The scoring method (0-1-2-3) was used to sum up the points to a total score ranging between 0 and 36, with a higher score indicating poorer mental health.

##### 2. Anxiety

The Arabic version of Taylor Manifest Anxiety Scale (TMAS) (Fahmi & Ghali, 1997) was used to separate normal participants from those with pathological levels of anxiety. It consisted of 50 true or false questions where an individual answers by reflecting on himself, to determine his/her anxiety level. The true response scores 1 point, so the total score ranges from 0 to 50. Scores from 0-16 denote no anxiety; 17-20 denote mild anxiety; 21-26 denote moderate anxiety; and 27-50 denote severe anxiety.

##### 3. Depression

The Arabic version of Beck Depression Inventory-II (BDI-II) (Ghareeb, 2000) was used as it is the most widely used instrument for detecting depression. It is a 21-question multiple-choice self-report inventory with four-point scale (0 to 3) for each item. Total score of 0 - 13 was considered minimal range, 14 - 19 was mild, 20 - 28 was moderate, and 29 - 63 was severe.

#### 2.2.2. Independent variables

##### Coping styles

It was assessed by the Brief-COPE scale, Arabic version (Nawel & Elisabeth, 2015). It is an abbreviated version of the COPE (Coping Orientation to Problems Experienced) Inventory. It is a self-report questionnaire designed to measure effective and ineffective ways to cope with a stressful life event. It is one of the best validated and most frequently used measures of coping strategies. The instrument consists of 28 items that measure 14 factors of 2 items each, which correspond to a Likert scale ranged from 0 – 3. Total scores on each scale range from 2 (minimum) to 8 (maximum). Higher scores indicate increased utilization of that specific coping strategy. Scores are presented for the two coping styles: (1) avoidant coping, which is characterized by the subscales of denial, substance use, venting, behavioral disengagement, self-distraction, and self-blame. Avoidant coping is associated with poorer physical health among those with CMI; and (2) approach coping is characterized by the subscales of active coping, positive reframing, planning, acceptance, seeking emotional support, and seeking informational support. Approach coping is associated with more helpful responses to adversity, including adaptive practical adjustment, better physical health outcomes and more stable emotional responding. Humor and religion items are neither categorized as approach or avoidant strategies.

### 2.3. Covariates

The following covariates were measured. Demographic variables included age, gender, education (no or primary education, secondary and university education), marriage (single, married, divorced, and widow) and the place of isolation (home and hospital). The severity of COVID-19 was categorized into mild, moderate, and severe cases according to the protocol of the Egyptian Ministry of Health and Population for diagnosis and treatment of COVID-19. Mild cases are the symptomatic cases with lymphopenia or leucopenia with no radiological lung affection by pneumonia. Moderate cases are defined as symptomatic patients with radiological features of pneumonia with or without leucopenia and lymphopenia. Severe cases are determined by the presence of any of the following: respiratory rate: > 30 / minute, SaO_2_ < 92 at room air, PaO_2_/FiO_2_ ratio < 300, chest radiology showing > 50% lung affection or progressive lung affection within 24 - 48 hours (Ministry of Health and Population, 2020).

### 2.4. Statistical Analyses

Data was entered and statistically analyzed on the Statistical Package of Social Science Software program, version 25(IBM SPSS Statistics for Windows). Data was presented as, mean, standard deviation, median and percentiles or quantitative variables and frequency and percentage for qualitative ones. Comparison between groups for qualitative variables was performed using Chi square or Fisher’s exact tests while for quantitative variables the comparison was conducted using independent sample t-test or Mann Whitney test. Correlation between numerical variables was presented. P values less than or equal to 0.05 were considered statistically significant.

## 3. Results

The total number of participants who completed the study questionnaire was 199 patients from the included quarantine centers. Patients were divided according to the presence of CMI into 2 groups: patients with COVID-19 and CMI (93 patients (46.73%)) and patients with COVID-19 without underlying CMI (106 patients (53.27%)). The demographic features of the studied groups are represented in **Table 1**. All patients did not have any previous psychiatric conditions. The chronic medical illnesses reported by participants were diabetes mellitus (n=38, 19.1%), systemic hypertension (n=40, 20.1%), bronchial asthma (n=20, 10.1%), ischemic heart disease (n=15, 7.5%) and untreated chronic hepatitis C infection (n=6, 3.0%).

**Table 1:**
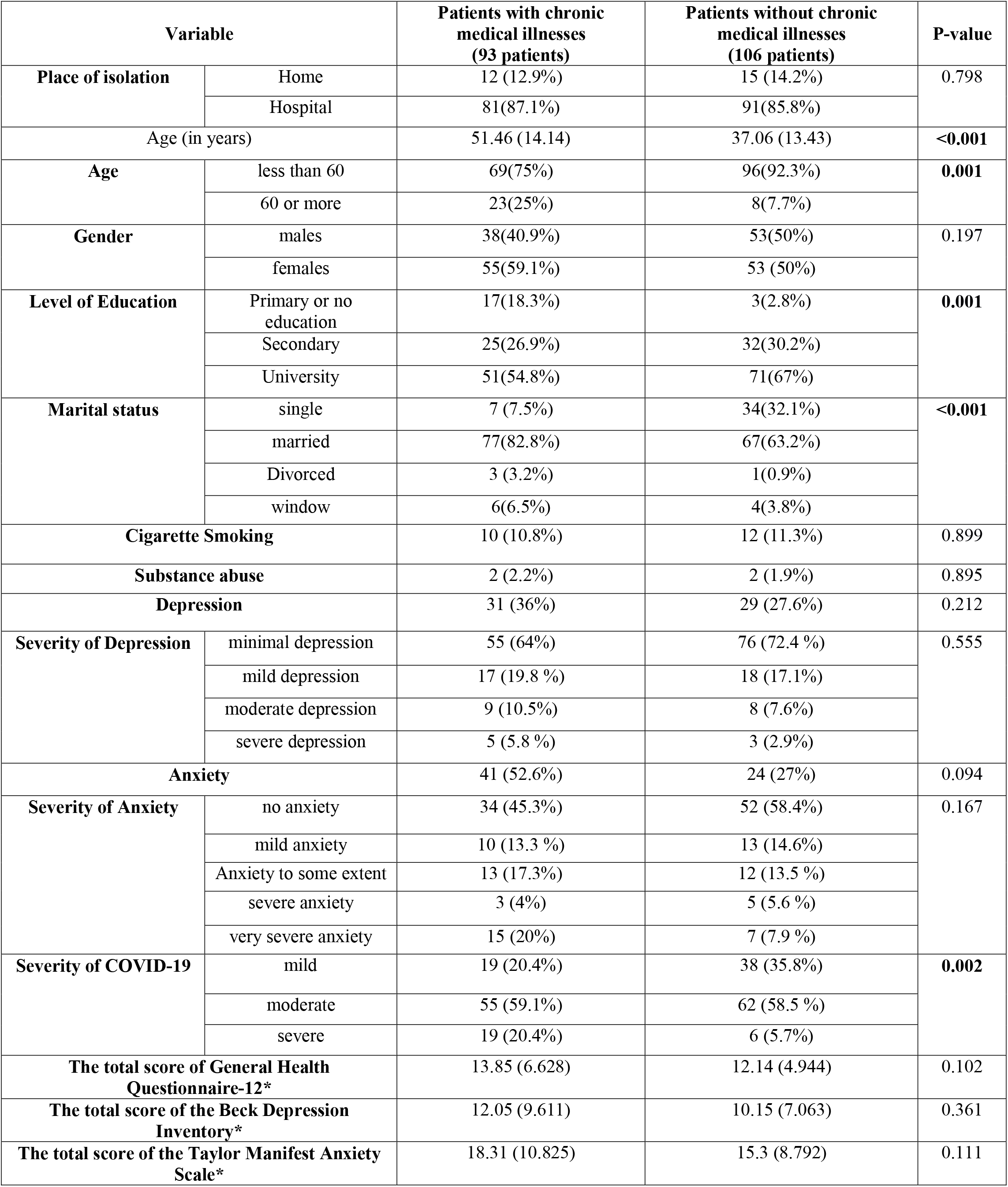
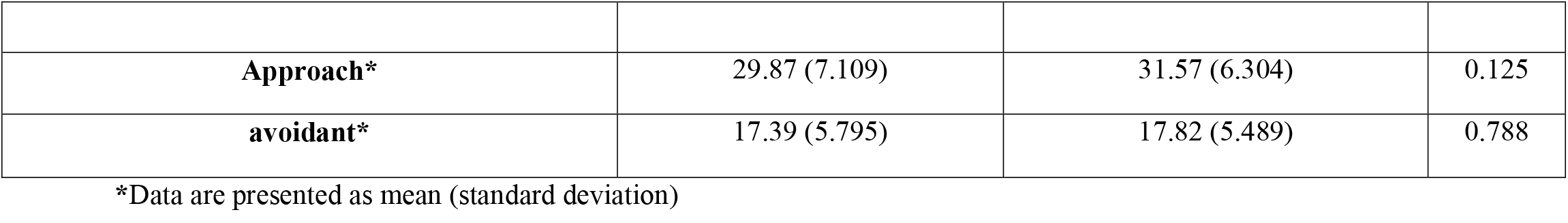
Demographic features of the studied cohort.

| Variable |  | Patients with chronic medical illnesses (93 patients) | Patients without chronic medical illnesses (106 patients) | P-value |
| --- | --- | --- | --- | --- |
| Place of isolation | Home | 12 (12.9%) | 15 (14.2%) | 0.798 |
|  | Hospital | 81(87.1%) | 91(85.8%) |  |
| Age (in years) |  | 51.46 (14.14) | 37.06 (13.43) | <0.001 |
| Age | less than 60 | 69(75%) | 96(92.3%) | 0.001 |
|  | 60 or more | 23(25%) | 8(7.7%) |  |
| Gender | males | 38(40.9%) | 53(50%) | 0.197 |
|  | females | 55(59.1%) | 53 (50%) |  |
| Level of Education | Primary or no education | 17(18.3%) | 3(2.8%) | 0.001 |
|  | Secondary | 25(26.9%) | 32(30.2%) |  |
|  | University | 51(54.8%) | 71(67%) |  |
| Marital status | single | 7 (7.5%) | 34(32.1%) | <0.001 |
|  | married | 77(82.8%) | 67(63.2%) |  |
|  | Divorced | 3 (3.2%) | 1(0.9%) |  |
|  | window | 6(6.5%) | 4(3.8%) |  |
| Cigarette Smoking |  | 10 (10.8%) | 12 (11.3%) | 0.899 |
| Substance abuse |  | 2 (2.2%) | 2 (1.9%) | 0.895 |
| Depression |  | 31 (36%) | 29 (27.6%) | 0.212 |
| Severity of Depression | minimal depression | 55 (64%) | 76 (72.4 %) | 0.555 |
|  | mild depression | 17 (19.8 %) | 18 (17.1%) |  |
|  | moderate depression | 9 (10.5%) | 8 (7.6%) |  |
|  | severe depression | 5 (5.8 %) | 3 (2.9%) |  |
| Anxiety |  | 41 (52.6%) | 24 (27%) | 0.094 |
| Severity of Anxiety | no anxiety | 34 (45.3%) | 52 (58.4%) | 0.167 |
|  | mild anxiety | 10 (13.3 %) | 13 (14.6%) |  |
|  | Anxiety to some extent | 13 (17.3%) | 12 (13.5 %) |  |
|  | severe anxiety | 3 (4%) | 5 (5.6 %) |  |
|  | very severe anxiety | 15 (20%) | 7 (7.9 %) |  |
| Severity of COVID-19 | mild | 19 (20.4%) | 38 (35.8%) | 0.002 |
|  | moderate | 55 (59.1%) | 62 (58.5 %) |  |
|  | severe | 19 (20.4%) | 6 (5.7%) |  |
| The total score of General Health Questionnaire-12* |  | 13.85 (6.628) | 12.14 (4.944) | 0.102 |
| The total score of the Beck Depression Inventory* |  | 12.05 (9.611) | 10.15 (7.063) | 0.361 |
| The total score of the Taylor Manifest Anxiety Scale* |  | 18.31 (10.825) | 15.3 (8.792) | 0.111 |

|  |  |  |  |
| --- | --- | --- | --- |
| **Approach\*** | 29.87 (7.109) | 31.57 (6.304) | 0.125 |
| **avoidant\*** | 17.39 (5.795) | 17.82 (5.489) | 0.788 |

\*Data are presented as mean (standard deviation)

Patients with underlying CMI were significantly older and had more severe COVID-19 disease presentations. There was no difference between the two groups regarding gender distribution, place of isolation, the occurrence of anxiety or depression during COVID-19 disease or their severity. Forty-one (52.6%) patients with CMI versus 24 (27%) patients without CMI during their COVID-19 disease had anxiety. Thirty-one (36%) patients with CMI developed depression during their COVID-19 disease (5.8% of them had severe depression) versus 29 (27.6%) patients without CMI developed depression during their COVID-19 disease (2.9% of them had severe depression) **(Table 1)**. Religion, emotional support, use of informational support and acceptance items of the Brief-COPE scale were the most used coping strategies by the study sample.

We found no significant difference in the coping strategies between patients with and without CMI except for the use of informational support that was significantly used by patients without CMI **(Table 2)**. Further statistical analyses to study the coping strategies within each CMI were done **(Supplementary tables 1-5)**. Patients without systemic hypertension showed significant use of approach coping strategies. Patients with chronic pulmonary illnesses showed significant use of avoidant coping. Active coping was significantly used by patients with ischemic heart disease.

**Table 2:**
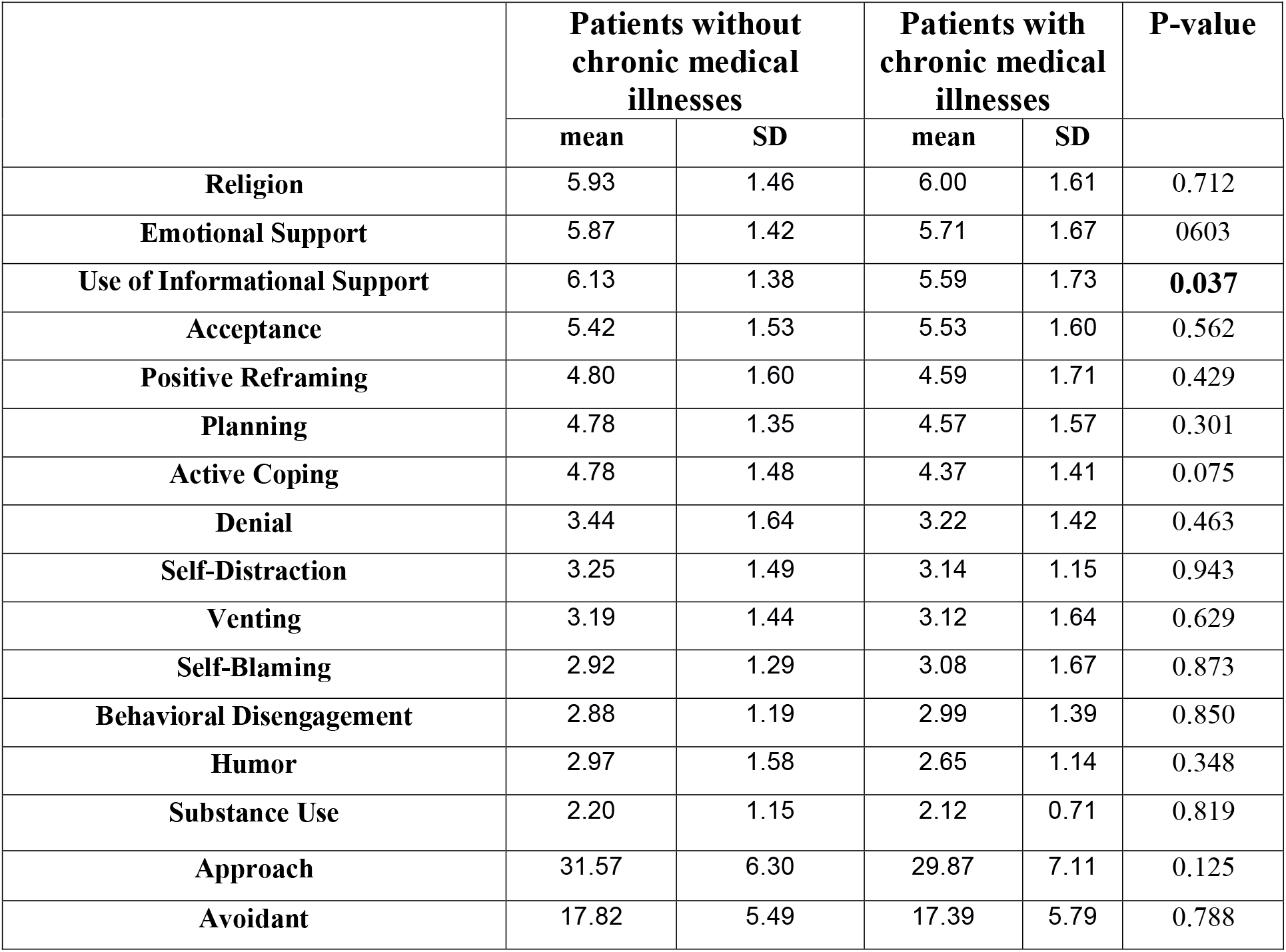
Coping strategies according to the presence of chronic medical illnesses.

|  | Patients without chronic medical illnesses |  | Patients with chronic medical illnesses |  | P-value |
| --- | --- | --- | --- | --- | --- |
|  | mean | SD | mean | SD |  |
| **Religion** | 5.93 | 1.46 | 6.00 | 1.61 | 0.712 |
| **Emotional Support** | 5.87 | 1.42 | 5.71 | 1.67 | 0.603 |
| **Use of Informational Support** | 6.13 | 1.38 | 5.59 | 1.73 | **0.037** |
| **Acceptance** | 5.42 | 1.53 | 5.53 | 1.60 | 0.562 |
| **Positive Reframing** | 4.80 | 1.60 | 4.59 | 1.71 | 0.429 |
| **Planning** | 4.78 | 1.35 | 4.57 | 1.57 | 0.301 |
| **Active Coping** | 4.78 | 1.48 | 4.37 | 1.41 | 0.075 |
| **Denial** | 3.44 | 1.64 | 3.22 | 1.42 | 0.463 |
| **Self-Distraction** | 3.25 | 1.49 | 3.14 | 1.15 | 0.943 |
| **Venting** | 3.19 | 1.44 | 3.12 | 1.64 | 0.629 |
| **Self-Blaming** | 2.92 | 1.29 | 3.08 | 1.67 | 0.873 |
| **Behavioral Disengagement** | 2.88 | 1.19 | 2.99 | 1.39 | 0.850 |
| **Humor** | 2.97 | 1.58 | 2.65 | 1.14 | 0.348 |
| **Substance Use** | 2.20 | 1.15 | 2.12 | 0.71 | 0.819 |
| **Approach** | 31.57 | 6.30 | 29.87 | 7.11 | 0.125 |
| **Avoidant** | 17.82 | 5.49 | 17.39 | 5.79 | 0.788 |

Regarding the whole study population, there were no gender differences in the coping strategies used by males and females (**Supplementary table 6)**. Young patients (below 60 years old) used approach strategies of active coping, positive reframing and planning significantly more than patients 60 years or older (**Supplementary table 7)**. Also, patients with university or higher education used active coping strategy more than patients with fewer education years (**Supplementary table 8)**. Avoidant coping strategies were significantly more used by divorced patients, home quarantined patients, patients who developed depression or anxiety during COVID-19 disease and patients who did not receive hydroxyl-chloroquine in their treatment regimens **(Supplementary tables 9-13)**. Regarding the severity of COVID-19 disease, approach coping strategies were more commonly used by patients with mild COVID-19 disease than patients with moderate and severe presentations **(Table 3)**.

**Table 3:** Coping strategies of the studied patients according to the severity of COVID-19 infection.

| Coping strategy | Mild COVID-19 | Moderate COVID-19 | Severe COVID-19 | P-value |
| --- | --- | --- | --- | --- |
| Religion | 6.25± 1.59 | 5.88± 1.53 | 5.72± 1.34 | 0.199 |
| Emotional Support | 6.12± 1.63 | 5.60± 1.52 | 5.96± 1.34 | 0.095 |
| Use of Informational Support Items | 6.55± 1.43 | 5.59± 1.55 | 5.76± 1.56 | **0.001** |
| Acceptance | 5.88± 1.66 | 5.36± 1.53 | 5.08± 1.35 | **0.038** |
| Positive Reframing | 5.06± 1.90 | 4.65± 1.60 | 4.20± 1.12 | 0.134 |
| Planning | 5.02± 1.71 | 4.63± 1.39 | 4.24± 0.97 | 0.135 |
| Active Coping | 5.02± 1.61 | 4.44± 1.36 | 4.36± 1.38 | 0.061 |
| Denial | 3.63± 1.73 | 3.32± 1.52 | 2.80± 1.08 | 0.117 |
| Self-Distracton | 3.52± 1.65 | 3.17± 1.23 | 2.68± 0.85 | 0.097 |
| Venting | 3.56± 1.74 | 3.00± 1.16 | 3.00± 2.20 | 0.114 |
| Self-Blaming | 3.10± 1.64 | 3.01± 1.46 | 2.72± 1.17 | 0.746 |
| Behavioral Disengagement | 3.00± 1.31 | 2.99± 1.34 | 2.52± 0.87 | 0.233 |
| Humor | 3.20± 1.83 | 2.74± 1.25 | 2.40± 0.71 | 0.373 |
| Substance Use | 2.08± 0.55 | 2.21± 1.19 | 2.16± 0.47 | 0.138 |
| Approach | 32.74± 8.55 | 30.10± 5.79 | 29.60± 5.12 | **0.011** |
| Avoidant | 18.49± 6.69 | 17.60± 5.28 | 15.88± 4.10 | 0.143 |

COVID-19 related anxiety correlated with denial (r= 0.272, P-value 0.001), behavioral disengagement (r= 0.258, P-value 0.001), venting (r= 0.293, P-value <0.001) and self-blaming (r= 0.260, P-value 0.001). The use of avoidant coping strategies showed significant positive correlation with COVID-19 related anxiety (r= 0.285, P-value <0.001). There was a significant negative correlation between the age of the patients with COVID-19 and depression, avoidant coping, and the use of humor. The total score of GHQ-12 showed highly significant positive correlation with depression (r= 0.573, P-value <0.001), anxiety (r= 0.491, P-value <0.001), Denial (r= 0.285, P-value <0.001), behavioral disengagement (r= 0.392, P-value <0.001), venting (r= 0.485, P-value <0.001), self-blaming (r= 0.442, P-value <0.001), the use of avoidant coping strategies (r= 0.447, P-value <0.001) and humor (r= 0.223, P-value 0.003). It also showed negative correlation with emotional support (r= -0.290, P-value <0.001) and the use of informational support (r= -0.166, P-value 0.026). There was a significant positive correlation between COVID-19 related depression and COVID-19 related anxiety disorders, use of informational support (r= 0.162, P-value 0.032) and venting strategies (r= 0.197, P-value 0.009) **(Table 4)**.

**Table 4:** Correlation between age, scores of anxiety, depression, and coping styles in the study sample.

|  |  | Age | The total score of General Health Questionnaire-12 | The total score of the Beck Depression Inventory | The total score of the Taylor Manifest Anxiety Scale | Approach Coping | Avoidant Coping |
| --- | --- | --- | --- | --- | --- | --- | --- |
| Age | Correlation coefficient | 1.000 | -0.096 | -.206** | -0.146 | -0.142 | -0.188* |
|  | P value |  | 0.179 | **0.005** | 0.062 | 0.057 | **0.012** |
| The total score of General Health Questionnaire-12 | Correlation coefficient | -0.096 | 1.000 | 0.573** | 0.491** | -0.104 | 0.477** |
|  | P value | 0.179 |  | **<0.001** | **<0.001** | 0.162 | **<0.001** |
| The total score of the Beck Depression Inventory | Correlation coefficient | -.206** | .573** | 1.000 | 0.684** | 0.088 | 0.102 |
|  | P value | **0.005** | **<0.001** |  | **<0.001** | 0.246 | 0.176 |
| The total score of the Taylor Manifest Anxiety Scale | Correlation coefficient | -0.146 | .491** | 0.684** | 1.000 | 0.046 | .285** |
|  | P value | 0.062 | **<0.001** | **<0.001** |  | 0.572 | **<0.001** |
| Self Distraction | Correlation coefficient | -.175* | .176* | -0.071 | 0.099 | .294** | .710** |
|  | P value | **0.019** | **0.017** | 0.348 | 0.216 | **<0.001** | **<0.001** |
| Active Coping | Correlation coefficient | -.202** | -0.026 | -0.019 | 0.011 | .603** | .235** |
|  | P value | **0.006** | 0.723 | 0.795 | 0.892 | **<0.001** | 0.001 |
| Denial | Correlation coefficient | -.149* | .285** | 0.091 | .272** | 0.083 | .726** |
|  | P value | **0.044** | **<0.001** | 0.223 | **0.001** | 0.265 | **<0.001** |
| Substance Use | Correlation coefficient | 0.034 | 0.118 | -0.121 | -0.083 | 0.038 | .284** |
|  | P value | 0.648 | 0.113 | 0.108 | 0.301 | 0.612 | **<0.001** |
| Emotional Support | Correlation coefficient | 0.098 | -.290** | -0.041 | -0.062 | .609** | -.204** |
|  | P value | 0.191 | **<0.001** | 0.586 | 0.443 | **<0.001** | 0.006 |
| Use of Informational Support | Correlation coefficient | -0.122 | -.166* | .162* | 0.067 | .623** | -.154* |
|  | P value | 0.105 | **0.026** | **0.032** | 0.410 | 0.000 | 0.039 |
| Behavioral Disengagement | Correlation coefficient | -0.098 | .392** | 0.044 | .258** | -.228** | .699** |
|  | P value | 0.189 | **<0.001** | 0.558 | **0.001** | 0.002 | **<0.001** |
| Venting | Correlation coefficient | -.206** | .485** | .197** | .293** | 0.111 | .749** |
|  | P value | **0.006** | **<0.001** | **0.009** | **<0.001** | 0.135 | **<0.001** |
| Positive Reframing | Correlation coefficient | -0.126 | -0.085 | 0.006 | 0.083 | .737** | .279** |
|  | P value | 0.092 | 0.251 | 0.936 | 0.302 | **<0.001** | **<0.001** |
| Planning | Correlation coefficient | -.214** | 0.036 | 0.086 | 0.007 | .728** | .317** |
|  | P value | **0.004** | 0.634 | 0.256 | 0.933 | **<0.001** | **<0.001** |
| Humor | Correlation coefficient | -.212** | .223** | 0.003 | 0.096 | .183* | .588** |
|  | P value | **0.005** | **0.003** | 0.968 | 0.234 | 0.014 | **<0.001** |
| **Acceptance** | Correlation coefficient | -0.075 | 0.010 | 0.061 | 0.089 | .691** | 0.127 |
|  | P value | 0.314 | 0.896 | 0.419 | 0.265 | 0.000 | 0.088 |
| **Religion** | Correlation coefficient | -0.015 | -0.043 | 0.115 | 0.060 | .548** | 0.135 |
|  | P value | 0.843 | 0.566 | 0.126 | 0.459 | 0.000 | 0.070 |
| **Self Blaming** | Correlation coefficient | -.183* | .442** | 0.123 | .260** | -0.065 | .736** |
|  | P value | **0.014** | **<0.001** | 0.102 | **0.001** | 0.388 | 0.000 |
| **Approach Coping** | Correlation coefficient | -0.142 | -0.104 | 0.088 | 0.046 | 1.000 | 0.152* |
|  | P value | 0.057 | 0.162 | 0.246 | 0.572 |  | **0.04** |
| **Avoidant Coping** | Correlation coefficient | -.188* | 0.477** | 0.102 | .285** | 0.152* | 1.000 |
|  | P value | **0.012** | **<0.001** | 0.176 | **<0.001** | **0.04** |  |

\* Correlation is significant at the 0.05 level (2-tailed).
\*\* Correlation is significant at the 0.01 level (2-tailed)

## 4. Discussion

The results of the present study emphasized on the difference in the scores of anxiety, depression and coping strategies between patients infected with COVID-19 disease with and without CMI during the peak of COVID-19 disease in Egypt (June-July 2020). Approach or adaptive coping strategies (emotional support, use of informational support and acceptance) and religion were more frequently used than avoidant or mal-adaptive strategies within the study sample, without statistical difference between patients with and without CMI (except for the use of information support item which was significantly used by patients without CMI). This showed that patients with COVID-19 disease tended to overcome their illness by focusing on the religious aspects, seeking emotional support and use of informational support to gain more knowledge about their condition and by accepting it. This might explain why there was no difference in the prevalence of anxiety and depression among patients with and without CMI as the use of several approach coping strategies might have increased the ability of the patients to adapt resulting into less anxiety and depressive symptoms and better psychological health.

**Umucu and Lee (2020)** have also found that the most frequent coping strategies among patients with COVID-19 and chronic conditions in the USA were acceptance and self-distraction, though their study included largely highly educated participants and they didn’t include a comparative group of COVID-19 patients without CMI. It was reported that the more the acceptance coping strategy used by patients with chronic illnesses, the better the quality of life (Elfström, Rydén, Kreuter, Taft, & Sullivan, 2005).

Denial was the most used avoidant coping strategy while substance use was least used by both patients with and without CMI in this study (without any statistical difference between them). This demonstrates that patients had a considerable tendency to deny the reality of the presence of the disease at some point. This was against the finding by **Umucu and Lee (2020)** who reported that denial was the least used coping strategy among their cohort. Generally, substance use and denial were previously demonstrated to be the least used coping strategies among patients with chronic medical diseases (Burker, Evon, Marroquin Loiselle, Finkel, & Mill, 2005; Tuncay, Musabak, Gok, & Kutlu, 2008). Research concerning denial as a coping strategy showed contradictory interpretations. Some studies reported denial as a negative adaptive mechanism for health and rehabilitation. **Umucu and Lee (2020)** have reported denial as a positive adaptive strategy that reduces stress in the short term and they suggested that denial could be of help for patients with COVID-19 and chronic conditions to draw their attention away from stress and negative emotional impacts. This could be supported by our study findings that denial negatively correlated with age and showed high significant positive correlation with COVID-19 related anxiety and psychological distress. Thus, it could be interpreted that young individuals and those who had psychiatric morbidities were liable to use denial to give them time to adjust to the distressing situation.

To our knowledge, this is the first research to study the different coping strategies among patients with the different COVID-19 disease severities. COVID-19 disease is classified by different guidelines into mild, moderate and severe categories or severe and non-severe categories. These different disease categories have different clinical presentations, levels of medical care, quarantine place and perception by the community (Epidemiology Working Group for NCIP Epidemic Response, 2020, Hao et al, 2020; Verity et al, 2020; WHO, 2020). Patients with mild COVID-19 were quarantined at home (according to the protocol of the Egyptian Ministry of Health and Population) [12], thus they had time and possibility to use informational support. In addition, their presence within their families facilitated the acceptance of their illness more than those with moderate and severe COVID-19 disease. This could explain why patients with mild COVID-19 disease had significant higher scores of approach coping than patients with moderate and severe COVID-19 disease.

Most of the published studies reported coping strategies in the general population or in medical and nursing staff and students rather than in patients with COVID-19 disease. **Skapinakis and colleagues (2020)** have reported about COVID-19 related coping strategies among the Greek population and found that acceptance, humor and planning were the most frequent used strategies. **Park and colleagues (2020)** found that self-distraction, active coping, and seeking emotional support were the most frequent used coping strategies in USA population (including Hispanic and non-Hispanic ethnic groups). **Yu and colleagues (2020)** found that patients with suspected COVID-19 disease within the Chinese population had less social support, spent long times seeking for information support items and rarely used any coping strategy to deal with their stressor. **Dawson and Golijani-Moghaddam (2020)** conducted a cross-sectional study in the UK where COVID-19 confirmed cases comprised 18% of their studied population and found that avoidant coping positively correlated with distress and negatively correlated with well-being.

As COVID-19 is a seriously evolving pandemic, this study may add the Egyptian experience to the global work of clinicians and researchers for a better understanding of coping strategies used by patients with COVID-19 disease across different cultures and communities.

## Conclusions

The current pandemic is a need for multidisciplinary research that should involve psychiatric and psychosocial aspects. Coping strategies to COVID-19 disease are different across populations. Understanding COVID-19 related stress and coping strategies in individuals with chronic medical conditions has implications for how individuals might be helped to manage their illness during the current presentation and intervene with the development of serious long-term mental health conditions in the future.

## Data Availability

The data that support the findings of this study are available on request from the corresponding author, [Shousha HI].

## Funding Statement

This research did not receive any specific grant from funding agencies in the public, commercial, or not-for-profit sectors.

## Acknowledgement

None

## Declarations of interest

None

## Ethical standards

The study was approved by the research ethics committee (N-36-2020 on 14/5/2020) of Faculty of Medicine, Cairo University and the research ethics committee of the Egyptian Ministry of Health and Population (15-2020/1 on 2/6/2020) and have therefore been performed in accordance with the ethical standards laid down in the 1964 Declaration of Helsinki and its later amendments. Details that might disclose the identity of the subjects under study were omitted.

## Data Accessibility statement

The data that support the findings of this study are available on request from the corresponding author. The data are not publicly available due to privacy of the patients.

## Notes

### Competing Interest Statement

The authors have declared no competing interest.

### Clinical Trial

NCT04459403

### Funding Statement

This research received no specific grant from any funding agency, commercial or not-for-profit sectors

### Author Declarations

The study was approved by the research ethics committee (N-36-2020 on 14/5/2020) of Faculty of Medicine, Cairo University and the research ethics committee of the Egyptian Ministry of Health and Population (15-2020/1 on 2/6/2020).

## References

Burker, E. J., Evon, D. M., Marroquin Loiselle, M., Finkel, J. B., Mill, M. R., 2005. Coping predicts depression and disability in heart transplant candidates. Journal of Psychosomatic Research, 59, 215–222. http://dx.doi.org/10.1016/j.jpsychores.2005.06.055v

Daradkeh, T. K., Ghubash, R., El-Rufaie, O. E. F., 2001. Reliability, Validity, and Factor Structure of the Arabic Version of the 12-Item General Health Questionnaire. Psychological Reports, 89(1), 85–94. doi:10.2466/pr0.2001.89.1.85

Dawson, D. L., Golijani-Moghaddam, N., 2020. COVID-19: Psychological flexibility, coping, mental health, and wellbeing in the UK during the pandemic. Journal of Contextual Behavioral Science, 17, 126–134. doi: 10.1016/j.jcbs.2020.07.010.

Elfström, M., Rydén, A., Kreuter, M., Taft, C., Sullivan, M., 2005. Relations between coping strategies and health-related quality of life in patients with spinal cord lesion. Journal of Rehabilitation Medicine, 37, 9–16. http://dx.doi.org/10.1080/16501970410034414

Epidemiology Working Group for NCIP Epidemic Response, Chinese Center for Disease Control and Prevention. [The epidemiological characteristics of an outbreak of 2019 novel coronavirus diseases (COVID-19) in China]. Zhonghua Liu Xing Bing Xue Za Zhi. 2020 Feb 10;41(2):145-151. Chinese. doi: 10.3760/cma.j.issn.0254-6450.2020.02.003. PMID: 32064853.

Fahmi, M., Ghali, M., 1997. Arabic version of Taylor Manifest Anxiety Scale. Egyptian Journal of Psychiatry, 11, 119–126.

Ghareeb, A., 2000. Manual of the Arabic BDI-II. Cairo, Egypt: Anglo Press.

Güler, A. A., Öztürk, M. A., 2020. COVID-19 in chronic diseases. Gazi Medical Journal - COVID 19 Special issue, 2(31), 266–270.

Hao, B., Sotudian, S., Wang, T., Xu, T., Hu, Y., Gaitanidis, A., Breen, K., Velmahos, G. C., Paschalidis, I. C., 2020. Early prediction of level-of-care requirements in patients with COVID-19. Elife 9, e60519. doi: 10.7554/eLife.60519.

Li, S., Wang, Y., Xue, J., Zhao, N., Zhu, T., 2020. The impact of COVID-19 epidemic declaration on psychological consequences: A study on active Weibo users. International Journal of Environmental Research and Public Health, 17, 2032. http://dx.doi.org/10.3390/ijerph17062032

Ministry of Health and Population, Egypt Management protocol for COVID-19 Patients Version 1.4 / 30th May 2020, Available at: https://www.elwatannews.com/data/iframe/pdf/17175200761591035127.pdf

Nawel, H.,Elisabeth. S., 2015. Adaptation and validation of the Tunisian version of the Brief COPE Scale. European Health Psychology, 17, 783.

Park, C. L., Russell, B. S., Fendrich, M., Finkelstein-Fox, L., Hutchison, M., Becker, J., 2020. Americans’ COVID-19 Stress, Coping, and Adherence to CDC Guidelines. Journal of General Internal Medicine, 35(8), 2296–2303. doi: 10.1007/s11606-020-05898-9. Epub 2020 May 29. PMID: 32472486; PMCID: PMC7259430.

Sanyaolu, A., Okorie, C., Marinkovic, A., Patidar, R., Younis, K., Desai, P., Hosein, Z., Padda, I., Mangat, J., Altaf, M., 2020. Comorbidity and its Impact on Patients with COVID-19. SN Comprehensive Clinical Medicine, 1–8. Advance online publication. https://doi.org/10.1007/s42399-020-00363-4

Skapinakis, P., Bellos, S., Oikonomou, A., Dimitriadis, G., Gkikas, P., Perdikari, E., Mavreas, V., 2020. Depression and Its Relationship with Coping Strategies and Illness Perceptions during the COVID-19 Lockdown in Greece: A Cross-Sectional Survey of the Population. Depression Research and Treatment, 3158954. doi: 10.1155/2020/3158954. PMID: 32908697; PMCID: PMC7450302.

Stanisławski, K., 2019. The Coping Circumplex Model: An Integrative Model of the Structure of Coping with Stress. Frontiers in Psychology, 10, 694. doi: 10.3389/fpsyg.2019.00694

Tuncay, T., Musabak, I., Gok, D. E., Kutlu, M., 2008. The relationship between anxiety, coping strategies and characteristics of patients with diabetes. Health and Quality of Life Outcomes, 6, 79. http://dx.doi.org/10.1186/1477-7525-6-79

Umucu, E., Lee, B., 2020. Examining the impact of COVID-19 on stress and coping strategies in individuals with disabilities and chronic conditions. Rehabilitation Psychology, 65(3), 193–198. doi: 10.1037/rep0000328.

Verity, R., Okell, L. C., Dorigatti, I., Winskill, P., Whittaker, C., Imai, N., Cuomo-Dannenburg, G., Thompson, H., Walker, P. G. T., Fu, H., Dighe, A., Griffin, J. T., Baguelin, M., Bhatia, S., Boonyasiri, A., Cori, A., Cucunubá, Z., FitzJohn, R., Gaythorpe, K., Green, W., Hamlet, A., Hinsley, W., Laydon, D., Nedjati-Gilani, G., Riley, S., van Elsland, S., Volz, E., Wang, H., Wang, Y., Xi, X., Donnelly, C. A., Ghani, A. C., Ferguson, N. M., 2020. Estimates of the severity of coronavirus disease 2019: a model-based analysis. The Lancet Infectious Diseases, 20(6), 669–677. doi: 10.1016/S1473-3099(20)30243-7. Epub 2020 Mar 30. Erratum in: Lancet Infect Dis. 2020 Apr 15: Erratum in: Lancet Infect Dis. 2020 May 4: PMID: 32240634; PMCID: PMC7158570.

WHO Report of the WHO-China Joint Mission on coronavirus disease 2019 (COVID-19). https://www.who.int/publications-detail/report-of-the-who-china-joint-mission-on-coronavirus-disease-2019-(covid-19). Date: Feb 28, 2020. Date accessed: October 18, 2020

Worldometers.info. 2020 October 8. Dover, Delaware, U.S.A. Retrieved from: https://www.worldometers.info/coronavirus/country/egypt/

Wu, Z., McGoogan, J. M., 2020. Characteristics of and important lessons from the coronavirus disease 2019 (COVID-19) outbreak in China: Summary of a Report of 72□314 Cases From the Chinese Center for Disease Control and Prevention. JAMA, 323:1239.

Yu, H., Li, M., Li, Z., Xiang, W., Yuan, Y., Liu, Y., Li, Z., Xiong, Z., 2020. Coping style, social support and psychological distress in the general Chinese population in the early stages of the COVID-19 epidemic. BMC Psychiatry, 20(1), 426. doi: 10.1186/s12888-020-02826-3. PMID: 32854656; PMCID: PMC7450895.

