## supplementary tables for "Psychiatric morbidities and Coping strategies in patients with different Coronavirus disease-2019 severities and chronic medical diseases: A multicenter cross-sectional study"

**Supplementary Table 1: Coping strategies in patients with and without Diabetes Mellitus**

| **Variables** | **Patients without Diabetes Mellitus** | | **Patients with Diabetes Mellitus** | | **P-value** |
| --- | --- | --- | --- | --- | --- |
|  | **mean** | **SD** | **mean** | **SD** |  |
| **Age** | 40.84 | 14.49 | 56.21 | 13.46 | **<0.001** |
| **Total General Health questionnaire** | 12.87 | 5.51 | 13.24 | 7.14 | 0.575 |
| **The total score of the Beck Depression Inventory** | 10.95 | 8.27 | 11.24 | 8.70 | 0.857 |
| **The total score of the Taylor Manifest Anxiety Scale** | 16.78 | 9.679 | 16.03 | 10.662 | 0.482 |
| **Religion** | 6.01 | 1.51 | 5.75 | 1.61 | 0.492 |
| **Emotional Support** | 5.78 | 1.53 | 5.88 | 1.57 | 0.752 |
| **Use of Informational Support** | 6.00 | 1.47 | 5.36 | 1.90 | 0.083 |
| **Acceptance** | 5.49 | 1.55 | 5.39 | 1.60 | 0.821 |
| **Positive Reframing** | 4.76 | 1.65 | 4.45 | 1.64 | 0.400 |
| **Planning** | 4.73 | 1.47 | 4.47 | 1.39 | 0.252 |
| **Active Coping** | 4.71 | 1.45 | 4.11 | 1.39 | **0.045** |
| **Denial** | 3.43 | 1.54 | 2.97 | 1.52 | **0.042** |
| **Self-Distraction** | 3.27 | 1.38 | 2.91 | 1.13 | 0.195 |
| **Venting** | 3.16 | 1.36 | 3.18 | 2.16 | 0.436 |
| **Self-Blaming** | 3.05 | 1.46 | 2.73 | 1.51 | 0.087 |
| **Behavioral Disengagement** | 2.97 | 1.27 | 2.73 | 1.33 | 0.191 |
| **Humor** | 2.91 | 1.50 | 2.42 | 0.75 | 0.193 |
| **Substance Use** | 2.16 | 0.96 | 2.18 | 1.03 | 0.671 |
| **Approach** | 31.05 | 6.74 | 29.64 | 6.60 | 0.175 |
| **Avoidant** | 17.83 | 5.49 | 16.70 | 6.19 | 0.145 |

**Supplementary Table 2: Coping strategies in patients with and without Systemic hypertension**

|  | **Patients without Hypertension** | | **Patients with Hypertension** | | **P-value** |
| --- | --- | --- | --- | --- | --- |
|  | **mean** | **SD** | **mean** | **SD** |  |
| **Age** | 40.99 | 14.93 | 54.85 | 12.63 | **<0.001** |
| **Total General Health questionnaire** | 12.70 | 5.11 | 13.90 | 8.12 | 0.919 |
| **The total score of the Beck Depression Inventory** | 11.01 | 7.98 | 10.97 | 9.76 | 0.579 |
| **The total score of the Taylor Manifest Anxiety Scale** | 16.23 | 9.593 | 18.62 | 10.864 | 0.336 |
| **Religion** | 6.03 | 1.47 | 5.69 | 1.73 | 0.302 |
| **Emotional Support** | 5.84 | 1.50 | 5.64 | 1.69 | 0.553 |
| **Use of Informational Support** | 6.01 | 1.48 | 5.39 | 1.84 | 0.057 |
| **Acceptance** | 5.55 | 1.51 | 5.14 | 1.71 | 0.145 |
| **Positive Reframing** | 4.78 | 1.62 | 4.42 | 1.78 | 0.212 |
| **Planning** | 4.82 | 1.48 | 4.14 | 1.22 | **0.015** |
| **Active Coping** | 4.72 | 1.45 | 4.08 | 1.40 | **0.018** |
| **Denial** | 3.38 | 1.55 | 3.18 | 1.54 | 0.397 |
| **Self-Distraction** | 3.22 | 1.39 | 3.11 | 1.14 | 0.874 |
| **Venting** | 3.16 | 1.36 | 3.17 | 2.09 | 0.448 |
| **Self-Blaming** | 2.99 | 1.42 | 3.03 | 1.70 | 0.742 |
| **Behavioral Disengagement** | 2.89 | 1.25 | 3.08 | 1.42 | 0.493 |
| **Humor** | 2.87 | 1.49 | 2.64 | 0.99 | 0.901 |
| **Substance Use** | 2.14 | 0.95 | 2.28 | 1.06 | **0.032** |
| **Approach** | 31.49 | 6.21 | 28.08 | 7.96 | **0.025** |
| **Avoidant** | 17.67 | 5.30 | 17.43 | 6.82 | 0.846 |

**Supplementary Table 3: Coping strategies in patients with and without pulmonary disorders**

|  | **Patients without pulmonary disorders** | | **Patients with pulmonary disorders** | | **P-value** |
| --- | --- | --- | --- | --- | --- |
|  | **mean** | **SD** | **mean** | **SD** |  |
| **Age** | 43.01 | 15.36 | 50.90 | 15.32 | **0.026** |
| **Total General Health questionnaire** | 12.50 | 5.79 | 16.90 | 4.83 | **<0.001** |
| **The total score of the Beck Depression Inventory** | 10.75 | 7.95 | 13.65 | 11.57 | 0.548 |
| **The total score of the Taylor Manifest Anxiety Scale** | 15.99 | 9.124 | 22.75 | 13.892 | 0.055 |
| **Religion** | 5.97 | 1.52 | 5.88 | 1.59 | 0.725 |
| **Emotional Support** | 5.86 | 1.51 | 5.19 | 1.72 | 0.118 |
| **Use of Informational Support** | 5.96 | 1.58 | 5.06 | 1.18 | **0.016** |
| **Acceptance** | 5.46 | 1.57 | 5.56 | 1.46 | 0.790 |
| **Positive Reframing** | 4.74 | 1.67 | 4.38 | 1.45 | 0.476 |
| **Planning** | 4.70 | 1.47 | 4.56 | 1.36 | 0.998 |
| **Active Coping** | 4.63 | 1.49 | 4.28 | 1.07 | 0.317 |
| **Denial** | 3.34 | 1.59 | 3.33 | 1.08 | 0.485 |
| **Self-Distraction** | 3.19 | 1.37 | 3.31 | 1.01 | 0.381 |
| **Venting** | 3.13 | 1.58 | 3.44 | 0.89 | 0.067 |
| **Self-Blaming** | 2.92 | 1.46 | 3.75 | 1.48 | **<0.001** |
| **Behavioral Disengagement** | 2.87 | 1.27 | 3.56 | 1.26 | **0.016** |
| **Humor** | 2.85 | 1.45 | 2.50 | 0.63 | 0.968 |
| **Approach** | 30.95 | 6.89 | 29.19 | 4.28 | 0.247 |
| **Avoidant** | 17.44 | 5.69 | 19.56 | 4.49 | **0.049** |

**Supplementary Table 4: Coping strategies in patients with Ischemic heart diseases**

|  | **Patients without Ischemic heart diseases** | | | **Patients with Ischemic heart diseases** | | | **P-value** |
| --- | --- | --- | --- | --- | --- | --- | --- |
|  | **Median** | **Percentile 25** | **Percentile 75** | **Median** | **Percentile 25** | **Percentile 75** |  |
| **Age** | 41.00 | 32.00 | 54.00 | 61.50 | 41.00 | 68.00 | **<0.001** |
| **Total General Health questionnaire** | 12.00 | 9.00 | 16.00 | 7.00 | 5.00 | 16.00 | 0.408 |
| **The total score of the Beck Depression Inventory** | 9.00 | 4.00 | 15.00 | 6.50 | 2.00 | 13.00 | 0.114 |
| **The total score of the Taylor Manifest Anxiety Scale** | 15.00 | 8.00 | 23.00 | 19.00 | 6.00 | 15.00 | 0.731 |
| **Religion** | 6.00 | 5.00 | 7.00 | 7.00 | 6.00 | 8.00 | 0.522 |
| **Emotional Support** | 6.00 | 4.00 | 7.00 | 6.00 | 4.00 | 8.00 | 0.315 |
| **Use of Informational Support** | 6.00 | 5.00 | 7.00 | 7.00 | 6.00 | 8.00 | 0.081 |
| **Acceptance** | 5.00 | 4.00 | 6.00 | 6.00 | 6.00 | 8.00 | 0.613 |
| **Positive Reframing** | 4.00 | 4.00 | 6.00 | 6.00 | 2.00 | 6.00 | 0.121 |
| **Planning** | 4.00 | 4.00 | 6.00 | 5.00 | 4.00 | 6.00 | 0.602 |
| **Active Coping** | 4.00 | 4.00 | 6.00 | 6.00 | 5.00 | 6.00 | **0.015** |
| **Denial** | 3.00 | 2.00 | 4.00 | 3.00 | 2.00 | 6.00 | 0.137 |
| **Self-Distraction** | 3.00 | 2.00 | 4.00 | 3.00 | 2.00 | 4.00 | 0.510 |
| **Venting** | 3.00 | 2.00 | 4.00 | 2.00 | 2.00 | 2.00 | 0.232 |
| **Self-Blaming** | 2.00 | 2.00 | 4.00 | 2.00 | 2.00 | 2.00 | 0.535 |
| **Behavioral Disengagement** | 2.00 | 2.00 | 4.00 | 2.00 | 2.00 | 2.00 | 0.544 |
| **Humor** | 2.00 | 2.00 | 3.00 | 2.00 | 2.00 | 3.00 | 0.209 |
| **Substance Use** | 2.00 | 2.00 | 2.00 | 2.00 | 2.00 | 2.00 | 0.052 |
| **Approach** | 30.00 | 26.00 | 35.00 | 38.00 | 28.00 | 39.00 | 0.078 |
| **Avoidant** | 16.00 | 13.00 | 21.00 | 15.00 | 14.00 | 17.00 | 0.806 |

**Supplementary Table 5: Coping strategies in patients with chronic hepatitis C**

|  | **Patients without chronic hepatitis C** | | **Patients with chronic hepatitis C** | | **P-value** |
| --- | --- | --- | --- | --- | --- |
|  | **Mean** | **SD** | **Mean** | **SD** |  |
| **Age** | 43.44 | 15.39 | 55.67 | 15.76 | 0.084 |
| **Total General Health questionnaire** | 13.06 | 5.81 | 9.17 | 6.01 | 0.116 |
| **The total score of the Beck Depression Inventory** | 11.08 | 8.37 | 7.50 | 6.56 | 0.372 |
| **The total score of the Taylor Manifest Anxiety Scale** | 16.72 | 9.859 | 13.50 | 9.434 | 0.544 |
| **Religion** | 5.94 | 1.52 | 6.60 | 1.67 | 0.333 |
| **Emotional Support** | 5.79 | 1.53 | 6.00 | 2.00 | 0.783 |
| **Use of Informational Support** | 5.88 | 1.55 | 6.20 | 2.49 | 0.395 |
| **Acceptance** | 5.43 | 1.55 | 6.80 | 1.10 | **0.043** |
| **Positive Reframing** | 4.72 | 1.64 | 4.40 | 2.19 | 0.906 |
| **Active Coping** | 4.57 | 1.46 | 5.20 | 1.30 | 0.234 |
| **Planning** | 4.68 | 1.44 | 5.00 | 2.24 | 0.652 |
| **Denial** | 3.33 | 1.53 | 3.80 | 2.05 | 0.677 |
| **Self-Distraction** | 3.21 | 1.35 | 3.00 | 1.00 | 0.889 |
| **Venting** | 3.18 | 1.54 | 2.40 | 0.89 | 0.171 |
| **Self-Blaming** | 3.01 | 1.48 | 2.40 | 0.89 | 0.336 |
| **Behavioral Disengagement** | 2.96 | 1.29 | 2.00 | 0.00 | 0.059 |
| **Humor** | 2.82 | 1.41 | 2.80 | 1.30 | 0.912 |
| **Substance Use** | 2.17 | 0.98 | 2.00 | 0.00 | 0.629 |
| **Approach** | 30.72 | 6.70 | 33.60 | 7.64 | 0.344 |
| **Avoidant** | 17.68 | 5.68 | 15.60 | 1.82 | 0.750 |

**Supplementary Table 6: Gender differences in coping strategies among patients with COVID-19 infection**

| **Variable** | **Males** | | **Females** | | **P-value** |
| --- | --- | --- | --- | --- | --- |
|  | **mean** | **SD** | **mean** | **SD** |  |
| **Total General Health questionnaire** | 11.74 | 4.93 | 13.95 | 6.35 | **0.021** |
| **Religion** | 5.99 | 1.44 | 5.94 | 1.60 | 0.792 |
| **Emotional Support** | 5.78 | 1.45 | 5.81 | 1.62 | 0.758 |
| **Use of Informational Support** | 5.79 | 1.60 | 5.97 | 1.55 | 0.491 |
| **Acceptance** | 5.37 | 1.62 | 5.56 | 1.50 | 0.362 |
| **Positive Reframing** | 4.53 | 1.67 | 4.86 | 1.63 | 0.176 |
| **Planning** | 4.75 | 1.27 | 4.63 | 1.61 | 0.370 |
| **Active Coping** | 4.68 | 1.40 | 4.51 | 1.51 | 0.404 |
| **Denial** | 3.32 | 1.50 | 3.36 | 1.59 | 0.995 |
| **Self-Distraction** | 3.25 | 1.37 | 3.16 | 1.32 | 0.705 |
| **Venting** | 3.10 | 1.27 | 3.21 | 1.74 | 0.904 |
| **Self-Blaming** | 3.01 | 1.55 | 2.98 | 1.41 | 0.845 |
| **Behavioral Disengagement** | 2.92 | 1.35 | 2.94 | 1.23 | 0.700 |
| **Humor** | 2.78 | 1.35 | 2.86 | 1.46 | 0.707 |
| **Substance Use** | 2.15 | 0.66 | 2.18 | 1.19 | 0.401 |
| **Approach** | 30.64 | 6.02 | 30.94 | 7.32 | 0.635 |
| **Avoidant** | 17.60 | 5.40 | 17.65 | 5.84 | 0.809 |

**Supplementary Table 7: Coping strategies of the studied patients according to age**

|  | **Less than 60 years** | | **60 years or above** | | **P-value** |
| --- | --- | --- | --- | --- | --- |
|  | **Mean** | **SD** | **Mean** | **SD** |  |
| **Total General Health questionnaire** | 13.21 | 5.98 | 11.45 | 5.15 | **NS** |
| **The total score of the Beck Depression Inventory** | 11.67 | 8.68 | 7.38 | 5.37 | **NS** |
| **Religion** | 5.99 | 1.50 | 5.81 | 1.67 | **NS** |
| **Emotional Support** | 5.75 | 1.56 | 6.04 | 1.43 | **NS** |
| **Use of Informational Support** | 5.93 | 1.57 | 5.62 | 1.55 | **NS** |
| **Acceptance** | 5.44 | 1.60 | 5.69 | 1.23 | **NS** |
| **Positive Reframing** | 4.80 | 1.62 | 4.08 | 1.70 | **0.039** |
| **Planning** | 4.76 | 1.48 | 4.12 | 1.07 | **0.041** |
| **Active Coping** | 4.73 | 1.42 | 3.86 | 1.55 | **0.004** |
| **Denial** | 3.42 | 1.56 | 2.93 | 1.46 | **NS** |
| **Self-Distraction** | 3.23 | 1.38 | 2.92 | 1.09 | **NS** |
| **Venting** | 3.19 | 1.59 | 2.77 | 0.95 | **NS** |
| **Self-Blaming** | 3.03 | 1.48 | 2.65 | 1.32 | **NS** |
| **Behavioral Disengagement** | 2.95 | 1.32 | 2.77 | 1.07 | **NS** |
| **Humor** | 2.87 | 1.45 | 2.50 | 1.10 | **NS** |
| **Substance Use** | 2.13 | 0.69 | 2.37 | 1.92 | **NS** |
| **Approach** | 30.99 | 6.83 | 29.42 | 5.72 | **NS** |
| **Avoidant** | 17.74 | 5.74 | 16.42 | 4.87 | **NS** |

**NS: non-significant, P-value>0.05**

**Supplementary Table 8: Coping strategies of the studied patients according to** **level of education**

|  | **Primary or No Education** | | **Secondary** | | **University or Higher** | | **P-value** |
| --- | --- | --- | --- | --- | --- | --- | --- |
|  | **Mean** | **SD** | **Mean** | **SD** | **Mean** | **SD** |  |
| **Age** | 61.15 | 15.22 | 37.50 | 32.00 | 40.50 | 32.00 | **<0.001** |
| **Total General Health questionnaire** | 13.90 | 7.56 | 13.00 | 9.00 | 12.00 | 9.00 | **NS** |
| **The total score of the Beck Depression Inventory** | 12.11 | 8.41 | 11.00 | 6.00 | 8.00 | 4.00 | **NS** |
| **Religion** | 6.19 | 1.17 | 6.32 | 1.59 | 5.76 | 1.51 | **NS** |
| **Emotional Support** | 6.06 | 1.44 | 5.51 | 1.56 | 5.89 | 1.53 | **NS** |
| **Use of Informational Support** | 6.06 | 1.44 | 5.85 | 1.39 | 5.88 | 1.68 | **NS** |
| **Acceptance** | 5.69 | 1.14 | 5.53 | 1.58 | 5.41 | 1.61 | **NS** |
| **Positive Reframing** | 4.50 | 1.46 | 4.83 | 1.60 | 4.68 | 1.71 | **NS** |
| **Planning** | 4.20 | 1.26 | 4.87 | 1.62 | 4.67 | 1.39 | **NS** |
| **Active Coping** | 3.50 | 1.25 | 4.72 | 1.49 | 4.70 | 1.41 | **0.002** |
| **Denial** | 2.94 | 1.55 | 3.36 | 1.73 | 3.39 | 1.45 | **NS** |
| **Self-Distraction** | 3.13 | 1.20 | 3.36 | 1.63 | 3.14 | 1.21 | **NS** |
| **Venting** | 2.69 | 0.87 | 3.34 | 1.69 | 3.14 | 1.52 | **NS** |
| **Self-Blaming** | 3.00 | 1.67 | 3.38 | 1.93 | 2.82 | 1.14 | **NS** |
| **Behavioral Disengagement** | 2.88 | 1.26 | 2.93 | 1.44 | 2.94 | 1.22 | **NS** |
| **Humor** | 2.06 | 0.25 | 3.17 | 1.56 | 2.77 | 1.38 | **0.004** |
| **Substance Use** | 2.00 | 0.00 | 2.30 | 1.59 | 2.12 | 0.58 | **NS** |
| **Approach** | 28.18 | 9.02 | 30.94 | 6.43 | 31.12 | 6.43 | **NS** |
| **Avoidant** | 15.76 | 6.63 | 18.62 | 6.95 | 17.43 | 4.65 | **NS** |

**NS: non-significant, P-value>0.05**

**Supplementary Table 9: Coping strategies of the studied patients according to marital status**

|  | **Single** | | **Married** | | **Divorced** | | **Widow** | | **P-value** |
| --- | --- | --- | --- | --- | --- | --- | --- | --- | --- |
|  | **Mean** | SD | **Mean** | SD | **Mean** | SD | **Mean** | SD |  |
| **Age** | 26.00 | 6.72 | 46.60 | 12.85 | 54.00 | 18.49 | 69.40 | 5.27 | **<0.001** |
| **Total General Health questionnaire** | 14.10 | 5.58 | 12.34 | 5.60 | 22.75 | 3.77 | 12.90 | 7.39 | **0.005** |
| **The total score of the Beck Depression Inventory** | 13.95 | 8.96 | 10.10 | 7.90 | 22.75 | 6.18 | 6.60 | 5.32 | NS |
| **Religion** | 6.03 | 1.72 | 5.98 | 1.49 | 5.00 | 1.41 | 5.70 | 1.25 | NS |
| **Emotional Support** | 5.32 | 1.53 | 5.90 | 1.51 | 2.00 | 1.41 | 6.60 | 1.07 | **0.020** |
| **Use of Informational Support** | 5.83 | 1.76 | 5.95 | 1.55 | 5.00 | 1.41 | 5.30 | 1.16 | NS |
| **Acceptance** | 5.68 | 1.63 | 5.39 | 1.58 | 6.00 | 1.41 | 5.70 | 0.95 | NS |
| **Positive Reframing** | 4.97 | 1.75 | 4.64 | 1.63 | 5.50 | 2.12 | 4.40 | 1.58 | NS |
| **Planning** | 5.05 | 1.43 | 4.62 | 1.46 | 5.50 | 2.12 | 4.00 | 1.05 | NS |
| **Active Coping** | 4.87 | 1.71 | 4.62 | 1.34 | 3.67 | 1.15 | 3.40 | 1.51 | **0.023** |
| **Denial** | 3.92 | 1.82 | 3.28 | 1.45 | 2.67 | 1.15 | 2.10 | 0.32 | **0.002** |
| **Self-Distraction** | 3.53 | 1.70 | 3.18 | 1.24 | 4.00 | 1.70 | 2.20 | 0.42 | **0.041** |
| **Venting** | 3.73 | 1.71 | 3.01 | 1.20 | 3.00 | 1.53 | 3.10 | 3.48 | **0.014** |
| **Self-Blaming** | 3.43 | 1.68 | 2.94 | 1.44 | 3.50 | 0.71 | 2.00 | 0.00 | **0.013** |
| **Behavioral Disengagement** | 3.32 | 1.36 | 2.85 | 1.27 | 3.50 | 2.12 | 2.40 | 0.70 | NS |
| **Humor** | 3.27 | 1.50 | 2.75 | 1.40 | 3.00 | 1.53 | 2.10 | 0.32 | **0.011** |
| **Substance Use** | 2.00 | 0.00 | 2.15 | 0.74 | 2.00 | 1.53 | 3.00 | 3.16 | 0.416 |
| **Approach** | 31.32 | 6.90 | 30.80 | 6.79 | 25.00 | 6.59 | 29.40 | 5.36 | NS |
| **Avoidant** | 19.82 | 6.40 | 17.19 | 5.31 | 21.00 | 6.10 | 14.80 | 4.39 | **0.017** |

**NS: non-significant, P-value>0.05**

**Supplementary Table 10: Coping strategies of the studied patients according to the place of quarantine**

|  | **Home quarantine** | | **Hospital quarantine** | | **P-value** |
| --- | --- | --- | --- | --- | --- |
|  | **Mean** | **SD** | **Mean** | **SD** |  |
| **Age** | 42.58 | 17.38 | 44.01 | 15.25 | NS |
| **Total General Health questionnaire** | 13.52 | 7.64 | 12.85 | 5.53 | NS |
| **The total score of the Beck Depression Inventory** | 14.08 | 9.86 | 10.52 | 8.00 | NS |
| **Religion** | 6.48 | 1.73 | 5.89 | 1.48 | NS |
| **Emotional Support** | 5.76 | 1.79 | 5.80 | 1.50 | NS |
| **Use of Informational Support** | 6.04 | 1.74 | 5.86 | 1.55 | NS |
| **Acceptance** | 6.30 | 1.64 | 5.35 | 1.51 | 0.011 |
| **Positive Reframing** | 5.25 | 1.98 | 4.63 | 1.59 | NS |
| **Planning** | 5.26 | 1.76 | 4.60 | 1.39 | NS |
| **Active Coping** | 5.08 | 1.68 | 4.52 | 1.41 | NS |
| **Denial** | 3.84 | 1.89 | 3.26 | 1.47 | NS |
| **Self-Distraction** | 3.63 | 1.50 | 3.14 | 1.31 | NS |
| **Venting** | 3.83 | 1.49 | 3.06 | 1.51 | 0.005 |
| **Self-Blaming** | 4.22 | 1.88 | 2.82 | 1.32 | <0.001 |
| **Behavioral Disengagement** | 3.42 | 1.25 | 2.86 | 1.28 | 0.012 |
| **Humor** | 3.48 | 1.75 | 2.73 | 1.32 | 0.018 |
| **Substance Use** | 2.16 | 0.80 | 2.16 | 1.00 | NS |
| **Approach** | 31.76 | 10.55 | 30.65 | 5.92 | NS |
| **Avoidant** | 20.08 | 7.49 | 17.23 | 5.19 | 0.016 |

**NS: non-significant, P-value>0.05**

**Supplementary Table 11: Coping strategies in patients with COVID-19 related depression**

|  | **Minimal depression** | | **Mild depression** | | **Moderate depression** | | **Severe depression** | | **P-value** |
| --- | --- | --- | --- | --- | --- | --- | --- | --- | --- |
|  | **Mean** | **SD** | **Mean** | **SD** | **Mean** | **SD** | **Mean** | **SD** |  |
| **Age** | 44.78 | 15.62 | 42.94 | 16.80 | 38.35 | 12.40 | 31.88 | 6.60 | NS |
| **Total GHQ** | 10.97 | 4.38 | 15.71 | 5.12 | 18.76 | 4.71 | 23.75 | 6.67 | NS |
| **Total Beck** | 6.48 | 3.66 | 16.14 | 1.68 | 24.06 | 2.73 | 34.88 | 5.30 | NS |
| **Religion** | 5.91 | 1.52 | 5.94 | 1.63 | 6.53 | 1.41 | 5.50 | 1.60 | NS |
| **Emotional Support** | 5.96 | 1.48 | 5.65 | 1.45 | 5.13 | 1.88 | 5.57 | 1.72 | NS |
| **Use of Informational Support** | 5.85 | 1.56 | 5.87 | 1.71 | 6.47 | 1.41 | 5.86 | 1.68 | NS |
| **Acceptance** | 5.54 | 1.52 | 5.16 | 1.85 | 5.33 | 1.29 | 6.50 | 1.20 | NS |
| **Positive Reframing** | 4.71 | 1.63 | 4.55 | 1.86 | 4.33 | 1.40 | 5.63 | 1.77 | NS |
| **Planning** | 4.65 | 1.35 | 4.53 | 1.48 | 5.13 | 1.81 | 5.25 | 2.19 | NS |
| **Active Coping** | 4.57 | 1.35 | 4.18 | 1.69 | 4.87 | 1.60 | 6.00 | 1.41 | **0.02** |
| **Denial** | 3.11 | 1.32 | 3.39 | 1.78 | 4.33 | 1.68 | 5.14 | 2.27 | **0.002** |
| **Self-Distraction** | 3.20 | 1.26 | 3.00 | 1.67 | 3.33 | 1.40 | 3.86 | 1.35 | NS |
| **Venting** | 2.93 | 1.41 | 3.45 | 1.79 | 3.80 | 1.47 | 4.71 | 1.60 | **0.001** |
| **Self-Blaming** | 2.77 | 1.22 | 3.00 | 1.81 | 3.73 | 1.79 | 4.75 | 1.75 | **0.001** |
| **Behavioral Disengagement** | 2.77 | 1.16 | 2.87 | 1.31 | 3.73 | 1.44 | 4.00 | 1.93 | **0.005** |
| **Humor** | 2.72 | 1.33 | 2.97 | 1.62 | 3.00 | 1.73 | 3.29 | 1.11 | NS |
| **Substance Use** | 2.18 | 1.04 | 2.00 | 0.00 | 2.00 | 0.00 | 2.86 | 2.27 | NS |
| **Approach** | 31.08 | 6.37 | 29.94 | 6.93 | 31.27 | 5.54 | 34.29 | 4.86 | NS |
| **Avoidant** | 16.84 | 4.41 | 17.81 | 6.47 | 20.93 | 5.59 | 25.71 | 9.66 | **0.002** |

**Supplementary Table 12: Coping strategies in patients with COVID-19 related anxiety**

|  | **No anxiety** | | **Mild anxiety** | | **Anxiety to some extent** | | **Severe anxiety** | | **Very severe anxiety** | | **P-value** |
| --- | --- | --- | --- | --- | --- | --- | --- | --- | --- | --- | --- |
|  | **Mean** | **SD** | **Mean** | **SD** | **Mean** | **SD** | **Mean** | **SD** | **Mean** | **SD** |  |
| **Age** | 44.16 | 15.02 | 42.43 | 15.56 | 42.83 | 18.34 | 41.25 | 14.25 | 38.41 | 15.82 | NS |
| **Total GHQ** | 10.67 | 4.77 | 13.13 | 4.54 | 13.96 | 3.75 | 16.75 | 3.45 | 20.68 | 7.43 | NS |
| **Religion** | 6.00 | 1.56 | 6.04 | 1.74 | 6.21 | 1.61 | 6.63 | 1.30 | 5.82 | 1.55 | NS |
| **Emotional Support** | 6.00 | 1.47 | 6.04 | 1.58 | 5.65 | 1.64 | 5.75 | 1.49 | 5.44 | 1.75 | NS |
| **Use of Informational Support** | 6.00 | 1.55 | 6.61 | 1.53 | 6.08 | 1.47 | 6.88 | 0.83 | 5.56 | 1.55 | NS |
| **Acceptance** | 5.53 | 1.56 | 5.70 | 1.74 | 5.54 | 1.64 | 4.88 | 1.13 | 6.35 | 1.17 | NS |
| **Positive Reframing** | 4.61 | 1.58 | 5.00 | 1.81 | 4.83 | 1.93 | 3.88 | 1.46 | 5.35 | 1.50 | NS |
| **Planning** | 4.73 | 1.32 | 4.55 | 1.74 | 5.00 | 1.62 | 3.75 | 1.04 | 5.29 | 1.96 | NS |
| **Active Coping** | 4.62 | 1.38 | 4.48 | 1.56 | 4.50 | 1.87 | 4.25 | 1.49 | 5.06 | 1.66 | NS |
| **Denial** | 2.92 | 1.20 | 3.22 | 1.57 | 3.87 | 1.66 | 4.75 | 1.49 | 4.44 | 2.23 | **0.0001** |
| **Self-Distraction** | 2.92 | 1.25 | 3.48 | 1.68 | 3.33 | 1.37 | 2.63 | 1.19 | 3.88 | 1.36 | **0.02** |
| **Venting** | 2.82 | 1.51 | 3.00 | 1.76 | 3.57 | 1.56 | 3.25 | 1.04 | 4.25 | 1.69 | **0.001** |
| **Self-Blaming** | 2.59 | 1.16 | 2.65 | 1.11 | 3.54 | 2.06 | 3.38 | 1.06 | 4.18 | 2.07 | **0.001** |
| **Behavioral Disengagement** | 2.55 | 0.92 | 2.52 | 0.95 | 3.00 | 1.57 | 3.13 | 0.99 | 4.29 | 1.93 | **0.0001** |
| **Humor** | 2.69 | 1.44 | 2.52 | 1.21 | 3.25 | 1.89 | 2.25 | 0.46 | 3.13 | 1.50 | NS |
| **Substance Use** | 2.24 | 1.24 | 2.00 | 0.00 | 2.00 | 0.00 | 2.00 | 0.00 | 2.38 | 1.50 | NS |
| **Approach** | 31.31 | 6.60 | 32.26 | 6.61 | 31.22 | 7.20 | 29.38 | 2.67 | 33.06 | 5.62 | NS |
| **Avoidant** | 15.99 | 4.22 | 16.87 | 5.43 | 18.87 | 5.32 | 19.13 | 4.45 | 23.88 | 8.37 | **0.0001** |

**Supplementary Table 13: Coping strategies of the studied patients according to hydroxyl-chloroquine intake**

|  | **No hydroxyl-chloroquine** | | | **Yes hydroxyl-chloroquine** | | | **P-value** |
| --- | --- | --- | --- | --- | --- | --- | --- |
|  | **Median** | **Percentile 25** | **Percentile 75** | **Median** | **Percentile 25** | **Percentile 75** |  |
| **Total General Health questionnaire** | 15.00 | 9.00 | 19.00 | 12.00 | 9.00 | 15.00 | **0.005** |
| **Self-Distraction** | 3.00 | 2.00 | 5.00 | 3.00 | 2.00 | 4.00 | **0.004** |
| **Active Coping** | 5.00 | 3.50 | 6.00 | 4.00 | 4.00 | 5.00 | 0.053 |
| **Denial** | 3.00 | 2.00 | 6.00 | 3.00 | 2.00 | 4.00 | **0.033** |
| **Substance Use** | 2.00 | 2.00 | 2.00 | 2.00 | 2.00 | 2.00 | 0.936 |
| **Emotional Support** | 5.00 | 4.00 | 7.00 | 6.00 | 5.00 | 7.00 | **0.002** |
| **Use of Informational Support** | 6.00 | 4.00 | 8.00 | 6.00 | 5.00 | 7.00 | 0.940 |
| **Behavioral Disengagement** | 3.00 | 2.00 | 4.00 | 2.00 | 2.00 | 4.00 | **0.01** |
| **Venting** | 3.00 | 3.00 | 4.00 | 2.00 | 2.00 | 4.00 | **<0.001** |
| **Positive Reframing** | 5.00 | 4.00 | 7.00 | 4.00 | 4.00 | 6.00 | 0.289 |
| **Planning** | 5.00 | 4.00 | 6.00 | 4.00 | 4.00 | 5.00 | **0.024** |
| **Humor** | 3.00 | 2.00 | 5.00 | 2.00 | 2.00 | 3.00 | **0.003** |
| **Acceptance** | 6.00 | 5.00 | 8.00 | 5.00 | 4.00 | 6.00 | **0.006** |
| **Religion** | 6.50 | 5.00 | 8.00 | 6.00 | 5.00 | 7.00 | **0.016** |
| **Self-Blaming** | 4.00 | 2.00 | 5.00 | 2.00 | 2.00 | 3.00 | **<0.001** |
| **Approach** | 32.50 | 25.00 | 38.00 | 30.00 | 27.00 | 35.00 | 0.485 |
| **Avoidant** | 19.50 | 16.00 | 24.00 | 15.00 | 13.00 | 21.00 | **<0.001** |
